# COVID-19 and acute kidney injury in German hospitals 2020

**DOI:** 10.1101/2021.04.30.21256331

**Authors:** Victor Walendy, Matthias Girndt, Daniel Greinert

## Abstract

**Introduction:** The SARS-CoV-2 pandemic is a major challenge for patients, healthcare professionals, and populations worldwide. While initial reporting focused mainly on lung involvement, the ongoing pandemic showed that multiple organs can be involved, and prognosis is largely influenced by multi-organ involvement. Our aim was to obtain nationwide retrospective population-based data on hospitalizations with COVID-19 and AKI.

**Materials & Methods:** We performed a query of G-DRG data for the year 2020 via the Institute for the hospital remuneration system (Institut für das Entgeltsystem im Krankenhaus GmbH, InEK) data portal and therefore included hospitalizations with a secondary diagnosis of RT-PCR proven COVID-19 infection, aged over 15 years. We included hospitalizations with acute kidney injury (AKI) stages 1 to 3. Age-specific and age-standardized hospitalization and in-hospital mortality rates (ASR) per 100.000 person years were calculated, with the German population of 2011 as the standard.

**Results:** In 2020, there were 16.776.845 hospitalizations in German hospitals. We detected 154.170 hospitalizations with RT-PCR proven COVID-19 diagnosis. The age-standardized hospitalization rate for COVID-19 in Germany was 232,8 per 100.000 person years (95% CI 231,6-233,9). The highest proportion of hospitalizations associated with COVID-19 were in the age group over 80 years. AKI was diagnosed in 16.773 (10,9%) of the hospitalizations with COVID-19. The relative risk of AKI for males was 1,49 (95%CI 1,44-1,53) compared to females. Renal replacement therapy (RRT) was performed in 3.443 hospitalizations, 20,5% of the hospitalizations with AKI. For all hospitalizations with COVID-19, the in-hospital mortality amounted to 19,7% (n= 30.300). The relative risk for in-hospital mortality was 3,87 (95%CI 3,80-3,94) when AKI occurred. The age-standardized hospitalization rates for COVID-19 took a bimodal course during the observation period. The first peak occurred in April (ASR 23,95 per 100.000 person years (95%CI 23,58-24,33)), hospitalizations peaked again in November 2020 (72,82 per 100.000 person years (95%CI 72,17-73,48)). The standardized rate ratios (SRR) for AKI and AKI-related mortality with the overall ASR for COVID-19 hospitalizations in the denominator, decreased throughout the observation period and remained lower in autumn than they were in spring. In contrast to all COVID-19 hospitalizations, the SRR for overall mortality in COVID-19 hospitalizations diverged from hospitalizations with AKI in autumn 2020.

**Discussion:** Our study for the first time provides nationwide data on COVID-19 related hospitalizations and acute kidney injury in Germany in 2020. AKI was a relevant complication and associated with high mortality. We observed a less pronounced increase in the ASR for AKI-related mortality during autumn 2020. The proportion of AKI-related mortality in comparison to the overall mortality decreased throughout the course of the pandemic.

## Introduction

The SARS-CoV-2 pandemic is a major challenge for patients, healthcare professionals, and populations worldwide. While initial reporting focused mainly on lung involvement, the ongoing pandemic showed that multiple organs can be involved, and prognosis is largely influenced by multi-organ involvement. According to published data, the kidney is the second most involved organ. In a study from the New York City area, 22,2% of hospitalized patients showed acute kidney injury (AKI)(1). A meta-analysis (2) confirmed the high AKI rate among COVID-19 patients. In contrast to this, there is also data with lower incidence of AKI(3,4) especially early analysis from Wuhan, China did not show high incidence rates of AKI (3–5). However, different definitions of AKI complicate comparability. In critically ill patients with COVID-19 as many as 55 to 90% experienced AKI (6–8), of whom 21 to 37% required renal replacement therapy (RRT) (6,8,9). The mortality rates of patients with acute kidney injury are significantly higher than in patients without AKI. In a study by Chan et al. (9), the in-hospital mortality was 50% among patients with, versus 8% among those without AKI. An analysis by Fisher et al. (8) found similar mortality rates (52% vs. 19.6%). In addition, the length of stay (LOS) was markedly prolonged when AKI occurred (9,10).

The German healthcare system offers comprehensive insurance coverage and, by international comparison, a high level of healthcare services. Throughout the COVID-19 pandemic there were no apparent acute care resource limitations, and the hospital capacities were at their limit but not overwhelmed. Nationwide unselected data, especially covering a longer time interval is not available. There is one observational study, which included 10.021 hospitalizations (11). The data from this study was obtained from one German health insurance company and covered a short period of time (from February 26 until April 19, 2020). In this study, AKI was not addressed.

Our aim was to obtain nationwide retrospective population-based data on hospitalizations with COVID-19 and AKI. Our query covered data from the year 2020 and contains, to our knowledge, the most comprehensive data regarding patients with COVID-19 and AKI in Germany.

## Materials & Methods

### Data Source

In accordance with the German Hospital Financing Act (KHG), a universal, performance-based remuneration system was introduced for general hospital services. The basis for this is the German-Diagnosis Related Groups system (G-DRG system), whereby each inpatient case of treatment is remunerated by means of a corresponding DRG rate. All hospitals submit their hospitalization data to the Institute for the hospital remuneration system (Institut für das Entgeltsystem im Krankenhaus GmbH, InEK). After submission, a plausibility control of the data is carried out by InEK. The submission of hospitalization data is mandatory for reimbursement of hospital stays. This leads to a strong incentive for hospitals to supply complete data, covering virtually every hospital in Germany. The transferred data include information on age, sex, discharge type, primary and secondary ICD-10-GM(12) coded diagnosis (International Classification of Diseases, 10th Edition, German Modification Version 2020) and performed operations and procedures (Operationen-und Prozedurenschlüssel Version 2020, OPS) (13). We performed a query of G-DRG data for the year 2020 via the InEK data portal (https://datenbrowser.inek.org/). The InEK data are available in aggregated form, so that differentiation by federal state is not possible. Differentiation by age can only be made in the predefined age groups and differentiation by gender only indirectly. The G-DRG data provided by InEK have been described in detail elsewhere (14).

### Cohort selection

We included hospitalizations with a secondary diagnosis of RT-PCR proven COVID-19 infection (ICD-10-GM: U07.1), aged over 15 years. We excluded hospitalizations 15.525 (9,1%) with hospital transfer and unknown discharge status from our analysis. We identified hospitalizations with acute kidney injury by their corresponding ICD-10-GM codes (S1 Table). We included hospitalizations with acute kidney injury stages 1 to 3 (AKIN 1 to 3) (15). We further searched for OPS codes indicating hemodialysis and mechanical ventilation (S1 Table). The survival status at the end of each hospitalization was obtained by discharge type. The observation period was from 01.02.2020 throughout 31.12.2020.

### Statistical analysis

We calculated crude, age-specific and age-standardized hospitalization rates (ASR) per 100.000 person years and the respective 95% confidence intervals (95% CI). We used the standard population of Germany in 2011 (16), provided by the Federal Bureau of Statistics (DESTATIS), for direct age-standardization (S2 Table). We further calculated standardized rate ratios (SRR), with their respective confidence intervals. Continuous variables are reported with standard deviation (SD). Categorical data is reported in absolute numbers and percentages. We used the ICD-10 version of the Elixhauser comorbidity index (ECI) to assess comorbidities (17). We excluded acute kidney injury as a comorbidity. All data analysis was carried out using R (Version 3.6.3) (18), for direct age-standardization the epitools package (19) was used, contingency tables were evaluated using the epiR package (20).

## Results

### General characteristics and age-standardized rates

In 2020, there were 16.776.845 hospitalizations in German hospitals. We detected 154.170 hospitalizations with RT-PCR proven COVID-19 diagnosis throughout the observation period. There were 79.781 (51,8%) male and 74.382 (48,2%) female patients hospitalized, respectively (Table 1). In 29.329 (19,0%) of the COVID-19 associated hospitalizations the treatment included an intensive care unit (ICU) stay. The age-standardized hospitalization rate for COVID-19 in Germany was 232,8 per 100.000 person years (95% CI 231,6-233,9) in 2020. Most hospitalizations (1394,1 per 100.000 person years) occurred in the age group of 80 years and older (Fig 1).

**Table 1.** General characteristics of COVID-19 hospitalizations in Germany 2020. ICU: intensive care unit - hospitalizations with intensive care treatment during the hospital stay, AKI: acute kidney injury - hospitalizations with acute kidney injury (Acute kidney injury network Stage 1-3), SD: standard deviation

|  | AKI |  | ICU |  | Overall |  |
| --- | --- | --- | --- | --- | --- | --- |
| N | 16.773 |  | 29.329 |  | 154.170 |  |
| Gender | % | n | % | n | % | n |
| male | 61,5 (10.308) |  | 62,6 (18.361) |  | 51,8 (79.781) |  |
| female | 38,5 (6.463) |  | 37,4 (10.965) |  | 48,2 (74.382) |  |
| Age group (years) | % | n | % | n | % | n |
| 16-17 | 0,0% | 1 | 0,1% | 38 | 0,3% | 472 |
| 18-29 | 0,4% | 70 | 1,7% | 493 | 4,0% | 6143 |
| 30-39 | 0,7% | 112 | 2,8% | 829 | 5,3% | 8185 |
| 40-49 | 1,9% | 326 | 5,8% | 1704 | 7,5% | 11601 |
| 50-54 | 2,6% | 443 | 5,8% | 1688 | 6,2% | 9537 |
| 55-59 | 4,5% | 754 | 7,9% | 2312 | 7,5% | 11602 |
| 60-64 | 6,8% | 1134 | 9,6% | 2818 | 7,7% | 11858 |
| 65-74 | 20,3% | 3408 | 21,8% | 6398 | 16,5% | 25418 |
| 75-79 | 17,0% | 2851 | 14,6% | 4282 | 11,7% | 18045 |
| 80+ | 45,8% | 7674 | 29,9% | 8767 | 33,3% | 51309 |
| length of stay (days, SD) | 18,4 (16,9) |  | 18,3 (16,7) |  | 11,3 (11,9) |  |
| Elixhauser comorbidity index (%<br>n) | % | n | % | n | % | n |
| 0 | 60 (10.041) |  | 65 (18.918) |  | 65 (99.783) |  |
| 1-4 | 24 (4.029) |  | 25 (7.186) |  | 25 (39.117) |  |
| >=5 | 16 (2.703) |  | 10 (3.225) |  | 10 (15.270) |  |
| mean (SD) | 2,0 (3,8) |  | 1,6 (3,8) |  | 1,2 (3,1) |  |

**Fig 1.**
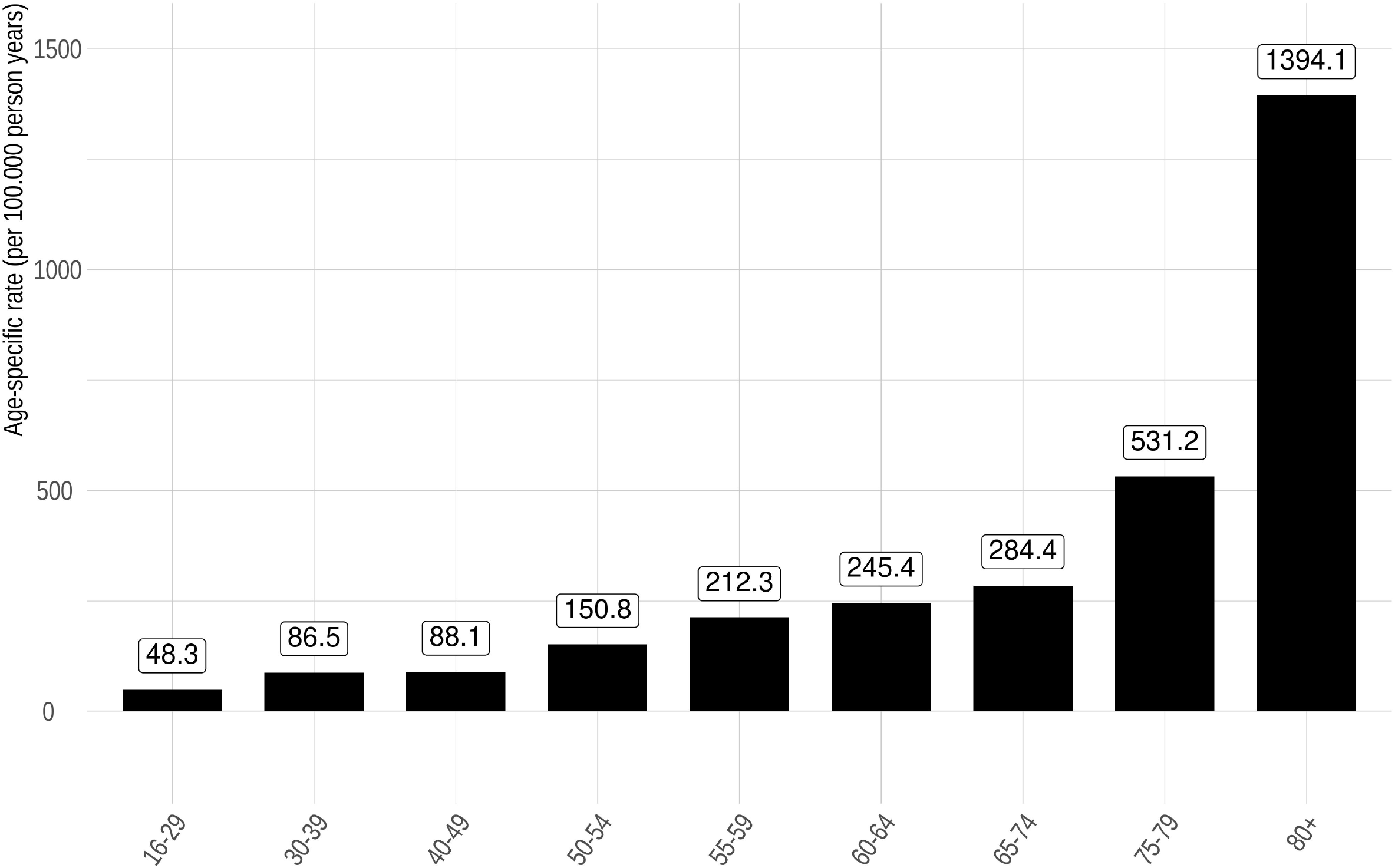
Age-specific rates of COVID-19 hospitalizations in Germany 2020 (per 100.000 person years)

### Hospital incidence of acute kidney injury

Acute kidney injury, including AKI Stages 1-3, was diagnosed in 16.773 (10,9%) of the hospitalized cases with COVID-19. The ASR for AKI was 25.8 (95%CI 25.4-26.2) per 100.000 person years (Table 2). While men and women were equally distributed among those hospitalized for COVID-19, men were more affected by AKI than women. The relative risk of AKI for males was 1,49 (95%CI 1,44-1,53) compared to females hospitalized with COVID-19 (S2 Table). We determined 10.310 (61,5%) male and 6.463 (38,5%) female hospitalizations with acute kidney injury, respectively (Table 1). The proportion of the severity of acute kidney injury was distributed as follows: 32,0% (n= 5.407) developed AKI Stage 1, AKI Stage 2 was present in 27,4 % (n= 4.625) and in 40,6% (n= 6.842) AKI Stage 3 was coded. There were 101 hospitalizations with more than one code for AKI. When acute kidney injury was present during hospital stay, 8.374 (49,9 %) needed intensive care. Furthermore, mechanical ventilation was required in 7.050 (42,0 %) hospitalizations with acute kidney injury (Table 2). The relative risk for hospitalizations with AKI to be treated in an intensive care unit was 3,27 (95%CI 3,21-3,34) (Table 3).

**Table 2:** Results COVID-19 hospitalizations in Germany 2020, ICU: intensive care unit - hospitalizations with intensive care treatment during the hospital stay, AKI: acute kidney injury - hospitalizations with acute kidney injury (acute kidney injury network, Stage 1-3), 95%CI: 95 % confidence interval, PYRs: person years

|  | AKI |  | ICU |  | Overall |  |
| --- | --- | --- | --- | --- | --- | --- |
|  | n | % | n | % | n | % |
| **Overall** | 16.773 (10,9%) |  | 29.329 (19,0%) |  | 154.170 (-) |  |
| **Renal replacement therapy** | 3.443 (20,5%) |  | 3.046 (10,4%) |  | - |  |
| **males** | 2.635 | 76,5% | 2.335 | 76,7% | - | - |
| **females** | 808 | 23,5% | 711 | 23,3% | - | - |
| **In-hospital mortality** | 9.717 (57,9%) |  | 12.322 (40,6%) |  | 30.300 (19,7%) |  |
| **males** | 6.283 | 64,7% | 7.990 | 64,8% | 17.635 | 58,2% |
| **females** | 3.434 | 35,3% | 4.330 | 35,2% | 12.665 | 41,8% |
| **AKI Stages** |  |  |  |  |  |  |
| **1** | 5.407 | 32,0% | 1.865 | 20,3% | - |  |
| **2** | 4.625 | 27,4% | 2.011 | 21,8% | - |  |
| **3** | 6.842 | 40,6% | 5.333 | 57,9% | - |  |
| **Chronic kidney disease** | 5.524 (32,9%) |  | 5.365 (18,3%) |  | 23.380 (15,2%) |  |
| **G3** | 3.638 | 21,7% | 3.225 | 11,0% | 15.270 | 9,9% |
| **G4** | 1.490 | 8,9% | 1.084 | 3,7% | 4.535 | 2,9% |
| **G5** | 396 | 2,4% | 1.056 | 3,6% | 3.575 | 2,3% |
| **Mechanical ventilation** | 7.050 (42,0%) |  | 17.144 (58,5%) |  | - |  |
| **males** | 5.052 | 71,6% | 11.508 | 67,1% | - |  |
| **females** | 1.998 | 28,3% | 5.634 | 32,9% | - |  |
| **Age-standardized rate (per 100.000 PYRs, 95% CI)** | 25,8 (25,4-26,2) |  | 44,0 (43,5-44,5) |  | 232,8 (231,6-233,9) |  |

**Table 3:** Contingency table for AKI and risk of ICU admission or in-hospital mortality, ICU: intensive care unit - hospitalizations with intensive care treatment during the hospital stay, AKI: acute kidney injury - hospitalizations with acute kidney injury (acute kidney injury network, Stage 1-3)

|  | ICU |  | Non-ICU |  | Overall |
| --- | --- | --- | --- | --- | --- |
|  | N | % | N | % | n |
| AKI | 8.374 | 49,9 | 8.399 | 50,1 | 16.773 |
| Non-AKI | 20.955 | 15,3% | 116.442 | 84,7% | 137.397 |
| Overall | 29.329 | 19,0% | 124.841 | 81,0% | 154.170 |
|  | Deceased |  | Alive |  | Overall |
|  | N | % | N | % | N |
| AKI | 9.717 | 57,9% | 7.056 | 42,1% | 16.773 |
| Non-AKI | 20.583 | 15,0% | 116.814 | 85,0% | 137.397 |
| Overall | 30.300 | 19,7% | 123.870 | 80,3% | 154.170 |

### Renal replacement therapy

When acute kidney injury was present, renal replacement therapy (RRT) was performed in 3.443 hospitalizations, 20,5% of hospitalizations with AKI and 2,2% of all COVID-19 associated hospitalizations. Furthermore, when RRT was conducted, mechanical ventilation was needed in 2.380 (69,1%) cases. The modality of renal replacement therapy was intermittent RRT in 230 (6,7%), and continuous or prolongend intermittend RRT in 3.213 (93,3%) hospitalizations, respectively. In 828 (24,0%) hospitalizations more than one procedure code indicating RRT was present.

Chronic kidney disease (CKD Stages 3 to 5) was present in 23.380 (15.2%) hospitalizations with AKI. We observed 3.638 (21,7%) hospitalizations with AKI and CKD Stage G3, in 1.490 (8.9%) hospitalizations CKD G4 was prevalent and 396 (2,4%) had CKD G5, respectively.

### In-hospital mortality

For all hospitalizations with COVID-19, the in-hospital mortality amounted to 19,7% (n= 30.300). Of these, 58,2% (n= 17.635) were men and 41,8% (n=12.665) were women (Table 2), respectively. The relative risk for in-hospital mortality for men amounted to 1,30 (95%CI 1,27-1,33) compared to women (S3 Table). The ASR for in-hospital mortality was 47,7 (95%CI 47,2-48,3) per 100.000 person years. Most deaths (511,2 per 100.000 person years) were observed in the age group over 80 years of age (Fig S1). When AKI was present, 9.717 died, accounting for 57,9% of deaths in this group (Table 2), the relative risk for in-hospital mortality was 3,87 (95%CI 3,80-3,94) when AKI occurred (Table 3). Moreover, when hemodialysis was performed, 2.891 died, which amounts to 61,4% of all hospitalizations with RRT.

### Length of stay

The mean length of stay (LOS) for hospitalizations with COVID-19 was 11,3 days (SD 11,9). When acute kidney injury was coded, the LOS increased to 18,4 days (SD 16,9). For cases that had an ICU stay the mean LOS was 18,3 days (SD 16,7) (Table 1).

### Time course of hospitalization rates and in-hospital mortality

The age-standardized hospitalization rates for COVID-19 took a bimodal course during the observation period. The first peak of ASR occurred in April 2020 (23,95 per 100.000 person years (95%CI 23,58-24,33)), after the ASR rose steadily beginning in February. Subsequently, ASR for COVID-19 hospitalizations peaked again in November 2020 (72,82 per 100.000 person years (95%CI 72,17-73,48)). In comparison to the first peak, ASR rose steadily for two months before reaching the highest rates (Fig 2A).

**Fig 2:**
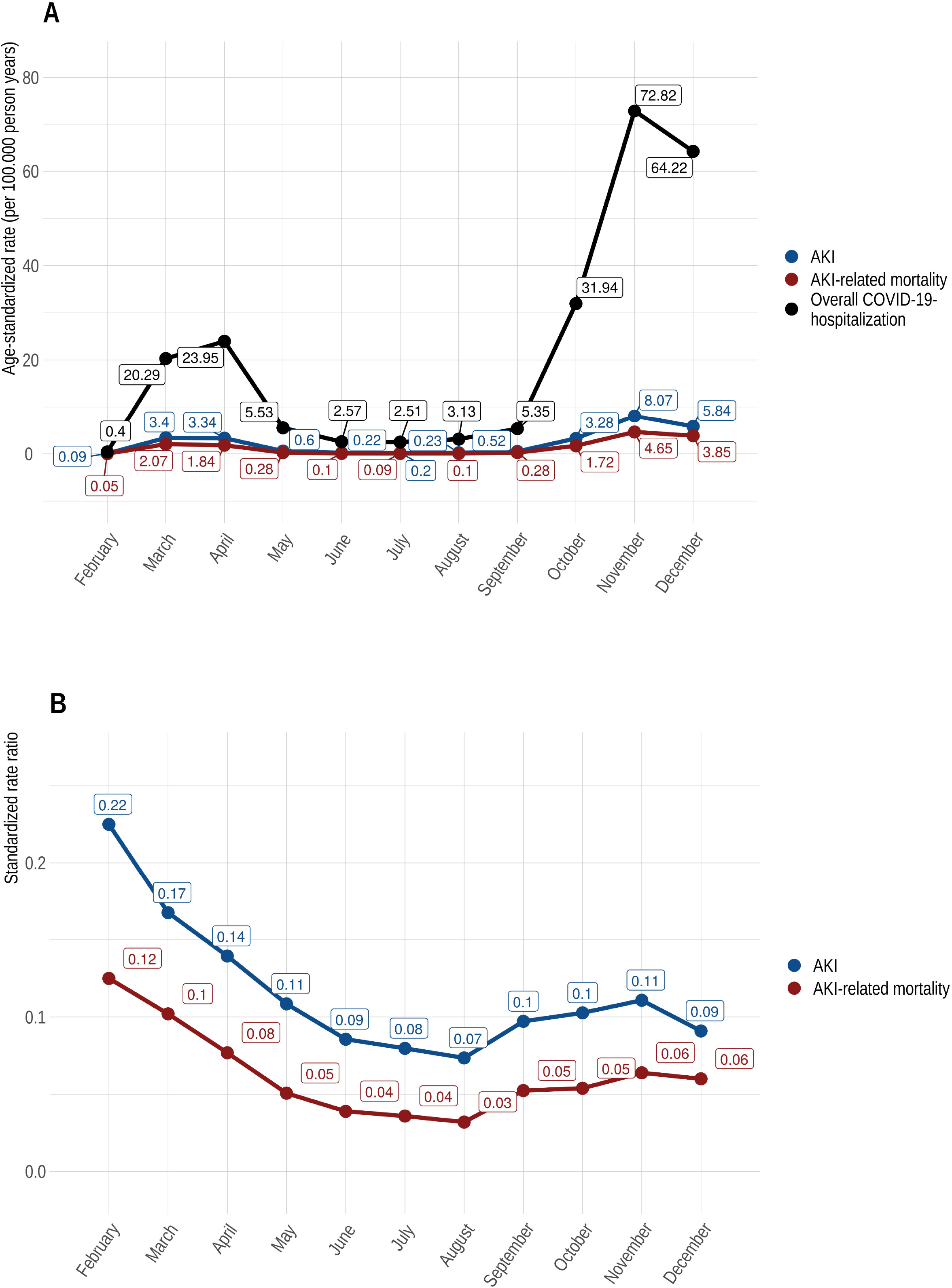
A) Time course of age-standardized rates (per 100.00 person years) of overall hospitalization, AKI and AKI-related mortality for COVID-19 in Germany (2020); B) Standardized rate ratios (SRR) of AKI age-standardized rates (per 100.00 person years) and AKI-related mortality in relation to the overall age-standardized COVID-19 hospitalization rate in Germany (2020)

The ASR for AKI and AKI-related mortality took a flatter curve, but overall follow a similar bimodal course. The ASR for AKI and AKI-related mortality reached their peak in March 2020 (for AKI 3,4 per 100.000 person years (95%CI 3,27-3,55), which was also when AKI-related mortality reached 2,07 (95%CI 1,96-2,18), respectively). In November 2020 ASR tipped again when ASR for AKI stood at 8,07 per 100.000 person years (95%CI 7,85-8,29), and the ASR for AKI-related mortality rose to 4,65 (95%CI 4,49-4,82)) (Fig 2A).

The standardized rate ratios (SRR) for AKI and AKI-related mortality with the overall ASR for COVID-19 hospitalizations in the denominator, actually decreased throughout the observation period (Fig 2B) and remained lower in autumn than they were in spring. In contrast to this finding, the ASR and in particular the SRR for overall mortality in COVID-19 hospitalizations rose sharply again as early as September 2020 (Fig 3A and 3B).

**Fig 3:**
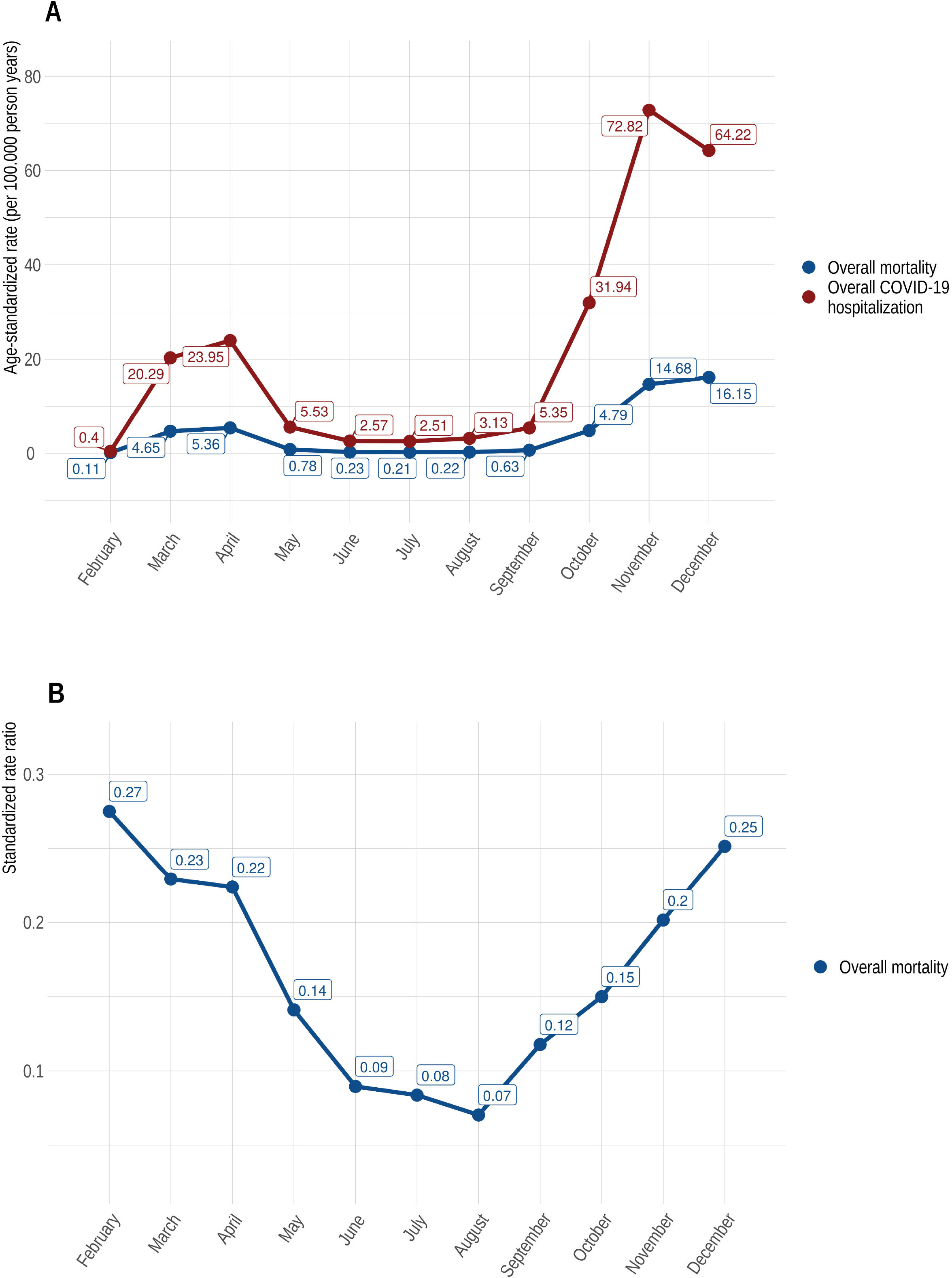
A) Time course of age-standardized rates (per 100.00 person years) of overall hospitalization and overall mortality for COVID-19 in Germany (2020); B) Standardized rate ratios (SRR) of overall mortality age-standardized rates (per 100.00 person years) in relation to the overall age-standardized COVID-19 hospitalization rate in Germany (2020)

The distribution of age groups varied throughout the observation period. In particular, the age group of 80 years and above were most prevalent in the months of April, November and December (Fig S4). Hospitalizations in which AKI was coded, were proportionately much older, with the 80 years and above age group being the most represented (Fig S5).

### Comorbidities

The Elixhauser comorbidity index (ECI) varied among the different groups. The mean ECI for all COVID-19 associated hospitalizations was 1,2 (SD 3,1). For hospitalizations requiring intensive care the mean ECI was 1,6 (SD 3,8). When acute kidney injury was present the ECI increased to 2,0 (SD 3,8). In general, the four most common coded comorbidities were uncomplicated hypertension (35,3%), fluid and electrolyte disorders (20,6%), uncomplicated diabetes mellitus (13,3%) and cardiac arrhythmias (10,0%) (Fig 4). When acute kidney injury or an ICU stay were coded, cardiac arrhythmias were relatively more prevalent compared to all hospitalizations with COVID-19 (Fig S2, S3).

**Fig 4:**
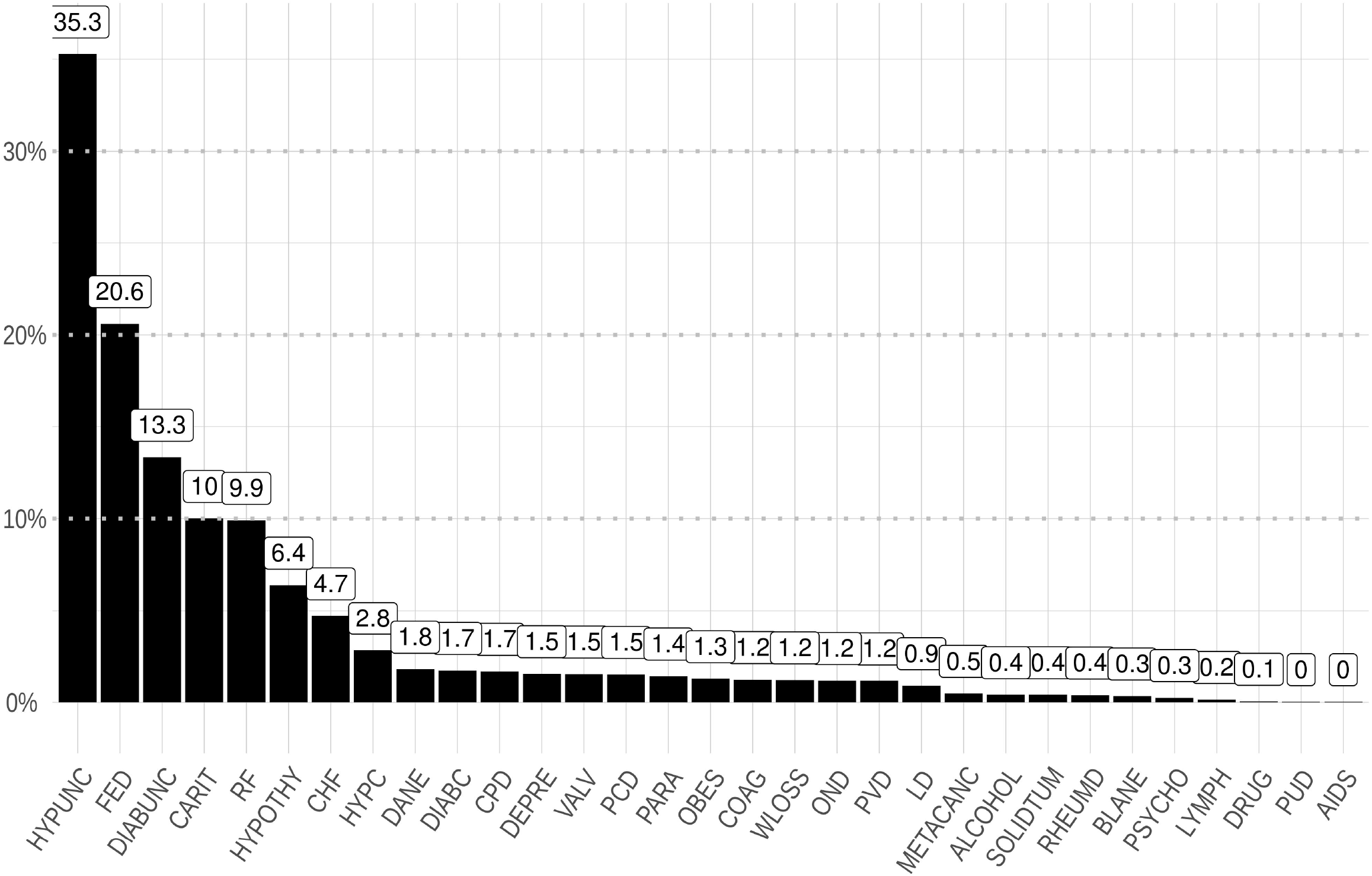
Relative frequencies of comorbidities in COVID-19 hospitalizations in Germany 2020. chf: congestive heart failure; carit: cardiac arrhythmias; valv: valvular disease; pcd: pulmonary circulation disorders; pvd: peripheral vascular disorders; hypunc: hypertension - uncomplicated; hypc: hypertension - complicated; para: paralysis; ond: other neurological disorders; cpd: chronic pulmonary disease; diabunc: diabetes -uncomplicated; diabc: diabetes-complicated; hypothy: hypothyroidism; ld: liver disease; pud: peptic ulcer disease-excluding bleeding; aids: AIDS/HIV; lymph: lymphoma; metacanc: metastatic cancer; solidtum: solid tumour: without metastasis; rheumd: rheumatoid arthritis/collaged vascular disease; coag: coagulopathy; obes: obesity; wloss: weight loss; fed: fluid and electrolyte disorders; blane: blood loss anaemia; dane: deficiency anaemia; alcohol: alcohol abuse; drug: drug abuse; psycho: psychoses; depre: depression;

## Discussion

Our study for the first time provides nationwide data on COVID-19 hospitalizations and acute kidney injury in Germany in 2020. The key findings that emerge from our data are as follows. The highest proportion of hospitalizations associated with COVID-19 were in the age group over 80 years. The gender distribution was equal in COVID-19 related hospitalization, however, we observed that men more often developed acute kidney injury (RR 1,49 (95%CI 1,44-1,53)), needed ICU care and had a higher risk of in-hospital mortality (RR: 1,30 (95%CI 1,27-1,33)) than women. Our results thus exhibit a gender bias. This is consistent with a meta-analysis including 3.111.714 COVID-19 associated hospitalizations worldwide, which identified male sex as a risk factor for death and ICU-admission, despite the gender ratio being equal at the time of admission (21). Different underlying mechanisms - from hormonal, genetic to behavioral effects and risk factor prevalence – have been discussed in the literature (21,22).

Overall, 19 % of all hospitalizations needed intensive care treatment. This fraction was particularly high in patients with acute kidney injury. This finding is in line with other studies (23,24). There are no clearly defined criteria for ICU admission in COVID-19 hospitalized patients. Germany possesses the highest capacity of intensive care beds in all OECD-countries. Despite the presumably higher availability of ICU capacity, this fact did not lead to higher utilization of these capacities. This could be an indicator that the percentage of critically ill patients was similar compared to different countries (23).

Occurrence of AKI considerably differs in previous studies, partly due to a different definition of AKI. Our analysis showed that 10.9% of all hospitalizations experienced acute kidney injury. The incidence of AKI in this national series is lower than what has been reported in regionally confined studies from the United States (8,9,23). A meta-analysis by Gabarre et al. (2) rendered similar results to our figures. They observed an average AKI incidence of 11% (95%CI 8–17%) overall, with highest ranges in the critically ill (23% (95%CI 14– 35%)). Nevertheless, the age structure, risk factor prevalence and the burden of COVID-19 during the observation period differ relevantly between countries and the AKI rate is therefore only comparable to a limited extend.

A large proportion of patients with acute kidney injury suffered severe kidney injury (AKIN Stage 3: 40,6%), this is consistent with previous findings (9). In the group of hospitalization with AKI 49,9% required ICU admission compared to 15,2 % of patients without AKI. The relative risk to be treated in an ICU was 3,27 for AKI patients. Moreover, in the group with AKI, 42,0 % required mechanical ventilation. Data from the early phase of the pandemic in New York state estimated that 53,6% of AKI patients needed invasive ventilation (10). This higher numbers are likely reflective of more severe disease outcomes in patients with AKI.

RRT was required in 2,2% of all hospitalizations. Among patients with AKI 20,5% required RRT. This figure is consistent with data from various studies (6,9,25). The majority (88,5%) of the RRT was performed in the intensive care unit. 69,1 % of all patients requiring dialysis needed invasive ventilation. Similar findings were reported in a multi-center cohort study from the United States (6). Previous studies suggested that there is an association between mechanical ventilation, AKI and RRT. RRT often occurs around time of intubation, suggesting a role for altered hemodynamics in this situation (6,10). Hence, vasopressors are often initiated around this time.

The overall in-hospital mortality for COVID-19 associated hospitalizations vary across different countries and cohorts, showing differences in testing and case identification, variable thresholds for hospitalization and staff capacities. Therefore, differences in in-hospital mortality ranged between 15 to 20% (26,27). Our analysis showed that 30.300 (19.7%) of hospitalizations with COVID-19 ultimately deceased, of 154.170 initially admitted to hospital. In-hospital mortality was 57.9% among patients with AKI versus 15% among those without AKI. This finding underlines the poor prognosis of these patients and the severity of disease.

The age-standardized hospitalization rate due to COVID-19 in 2020 was bimodally shaped, with peak rates in April and November. This observation is congruent with the incidence figures reported by the Robert Koch Institute (28). The first peak with increasing mortality rates in March and April was followed by a decline in mortality rates and a concomitant decrease in hospital admissions. Similar results have already been demonstrated in various studies from other countries. In a data-analysis from the United States a decrease in the risk-adjusted mortality was observed, ranging from 16.56 % to 9.29% in the early period of this study (January through April 2020) compared with the later period (May through June 2020) (29). Further, national ICU data from the United Kingdom – published as a preprint up to now - confirmed this finding. The authors found a reduction in mortality rates from 41,4% in March 2020 to 24,8% in June 2020 (30). The underlying reason for the decrease of mortality rates is not obvious and certainly multifactorial. One important factor was certainly the concomitant decline in the incidence rate in the UK and the US. For example, a large US study of health insurance data, involving more than 38.517 patients, revealed that the strongest determinant of improvements in in-hospital outcome was a decline in community rates of infection (29). Other reasons might be a shift in demographics, experience in physicians to early diagnose and treat COVID-19 patients, hospitals and staff became less overwhelmed during time. In our data during the second peak in COVID-19 hospitalizations, which was more pronounced than the first, the in-hospital mortality rate increased steadily from September to December and exceeded the COVID-19 associated in-hospital mortality in March and April markedly (5,36 per 100.000 person years in April and 16,15 per 100.000 person years in December). One can only speculate about the factors that led to this development. One, but certainly not the only, explanation could be a different age distribution of patients admitted to the hospital with a higher proportion of older patients in November and December, as in our data (Fig S4, Fig S5). Further, multiple factors might have influenced in-hospital mortality. The increasing number of patients in need of critical care could have overwhelmed hospitals regionally, which happened for example in Saxony, Germany in December 2020. As already mentioned, an American study has shown that hospitals did better when the prevalence of COVID-19 in their surrounding communities was lower(29). Another contributing factor could have been staff shortages driven by COVID-19 infections, school lockdowns, quarantine regulations and the necessity to take care of relatives infected with COVID-19 or in quarantine. In contrast to the development discussed above, the ASR for AKI and AKI-related mortality had a disproportionately smaller increase and were already declining in December. The age distribution in our cohort was similar among all hospitalized patients, with a higher proportion of elderly patients in the second peak in October, November and December in comparison to February, March and April. Thus, differences in age distribution is not a good explanation for this finding (S4 Fig and S5). At this point, it must be said again that multiple factors could have influenced the ASR for AKI und AKI-related mortality. Apart from differences in the patient population or age distribution, changes in the treatment strategy - e.g., the early use of drug therapies for patients with an increased risk of severe COVID-19 courses.

The mean length of stay (LOS-discharged or dead) of all patients was 11,3 days and increased with AKI to 18,4 days. LOS varies in different studies, and - according to a systematic review by Rees et al. (31) - the median LOS was 14 days in China, compared with 5 (IQR 3–9) days outside of China. Similar figures were reported by Fisher et al. (8). This may be explained by many differences in criteria for admission and discharge between countries, and different timings in the course of the pandemic. Hospital capacities in Germany were not overwhelmed by the pandemic, and the pressure to discharge patients might have been lower. The most comprehensive analysis of German patients to date included 10.021 patients with a mean LOS of 14,3 days (32). This was longer than in our analysis, an explanation for the discrepancy could be timing of data collection. Karagiannidis et al. collected their data at the beginning of the pandemic. Due to an increasing experience in treating COVID-19 patients and new treatment options, the length of stay consequently declined.

The observed comorbidities varied between the different groups. Hypertension, diabetes mellitus, fluid and electrolyte disorders and cardiac arrhythmias were the most commonly coded comorbidities. This finding is plausible, as these are the most common comorbidities in patients with chronic kidney disease (33) and likewise known risk factors for hospitalization with COVID-19 (34). Cardiac arrhythmias became more prevalent in hospitalizations with AKI. Cardiac arrhythmias are a common finding in COVID-19, in particular with an increasing number of comorbidities (35).

## Strengths and limitations

The major strength of this study is that we provide a nationwide population-based view on COVID-19 hospitalizations with acute kidney injury in Germany. We retrieved DRG-data covering virtually every hospital in Germany. In this study, we use routine hospitalization data, which is lacking clinical detail. This might lead to confounding that we could not adjust for. The InEK provides aggregated data due to data safety precautions, which is fully anonymized. It is, therefore, possible that this could lead to bias caused by readmission, though this possibility is limited, because of the severity of most cases and our efforts to exclude readmissions from the dataset. Another important fact to consider, when analyzing routine data, is the observation period. In our analysis the observation period ends in the midst of the second wave of the COVID-19 pandemic in Germany. This will lead to bias, as the outcome of these cases remains unknown, in particular for hospitalizations in the last month of the observation period.

## Data Availability

Data is fully accessible

https://datenbrowser.inek.org/

## Supporting information

S1 Fig. Age-specific mortality rates of COVID-19 hospitalizations in Germany 2020 (per 100.000 person years)

S2 Fig. Relative frequencies of comorbidities in COVID-19 hospitalizations with AKI in Germany 2020.

chf: congestive heart failure; carit: cardiac arrhythmias; valv: valvular disease; pcd: pulmonary circulation disorders; pvd: peripheral vascular disorders; hypunc: hypertension - uncomplicated; hypc: hypertension - complicated; para: paralysis; ond: other neurological disorders; cpd: chronic pulmonary disease; diabunc: diabetes -uncomplicated; diabc: diabetes- complicated; hypothy: hypothyroidism; ld: liver disease; pud: peptic ulcer disease-excluding bleeding; aids: AIDS/HIV; lymph: lymphoma; metacanc: metastatic cancer; solidtum: solid tumour: without metastasis; rheumd: rheumatoid arthritis/collaged vascular disease; coag: coagulopathy; obes: obesity; wloss: weight loss; fed: fluid and electrolyte disorders; blane: blood loss anaemia; dane: deficiency anaemia; alcohol: alcohol abuse; drug: drug abuse; psycho: psychoses; depre: depression;

S3 Fig. Relative frequencies of comorbidities in COVID-19 hospitalizations with ICU treatment in Germany 2020.

chf: congestive heart failure; carit: cardiac arrhythmias; valv: valvular disease; pcd: pulmonary circulation disorders; pvd: peripheral vascular disorders; hypunc: hypertension - uncomplicated; hypc: hypertension - complicated; para: paralysis; ond: other neurological disorders; cpd: chronic pulmonary disease; diabunc: diabetes -uncomplicated; diabc: diabetes-complicated; hypothy: hypothyroidism; ld: liver disease; pud: peptic ulcer disease-excluding bleeding; aids: AIDS/HIV; lymph: lymphoma; metacanc: metastatic cancer; solidtum: solid tumour: without metastasis; rheumd: rheumatoid arthritis/collaged vascular disease; coag: coagulopathy; obes: obesity; wloss: weight loss; fed: fluid and electrolyte disorders; blane: blood loss anaemia; dane: deficiency anaemia; alcohol: alcohol abuse; drug: drug abuse; psycho: psychoses; depre: depression;

S4 Fig. Percentage distribution of age groups during the observation period (Overall COVID-19)

S5 Fig. Percentage distribution of age groups during the observation period (AKI)

S1 Table. List of ICD-10-GM and OPS Codes, version 2020

S2 Table. Contingency table AKI by sex

S3 Table. Contingency table in-hospital mortality by sex

