## Supplementary figures and images for "COVID-19 and acute kidney injury in German hospitals 2020"

### Fig S1

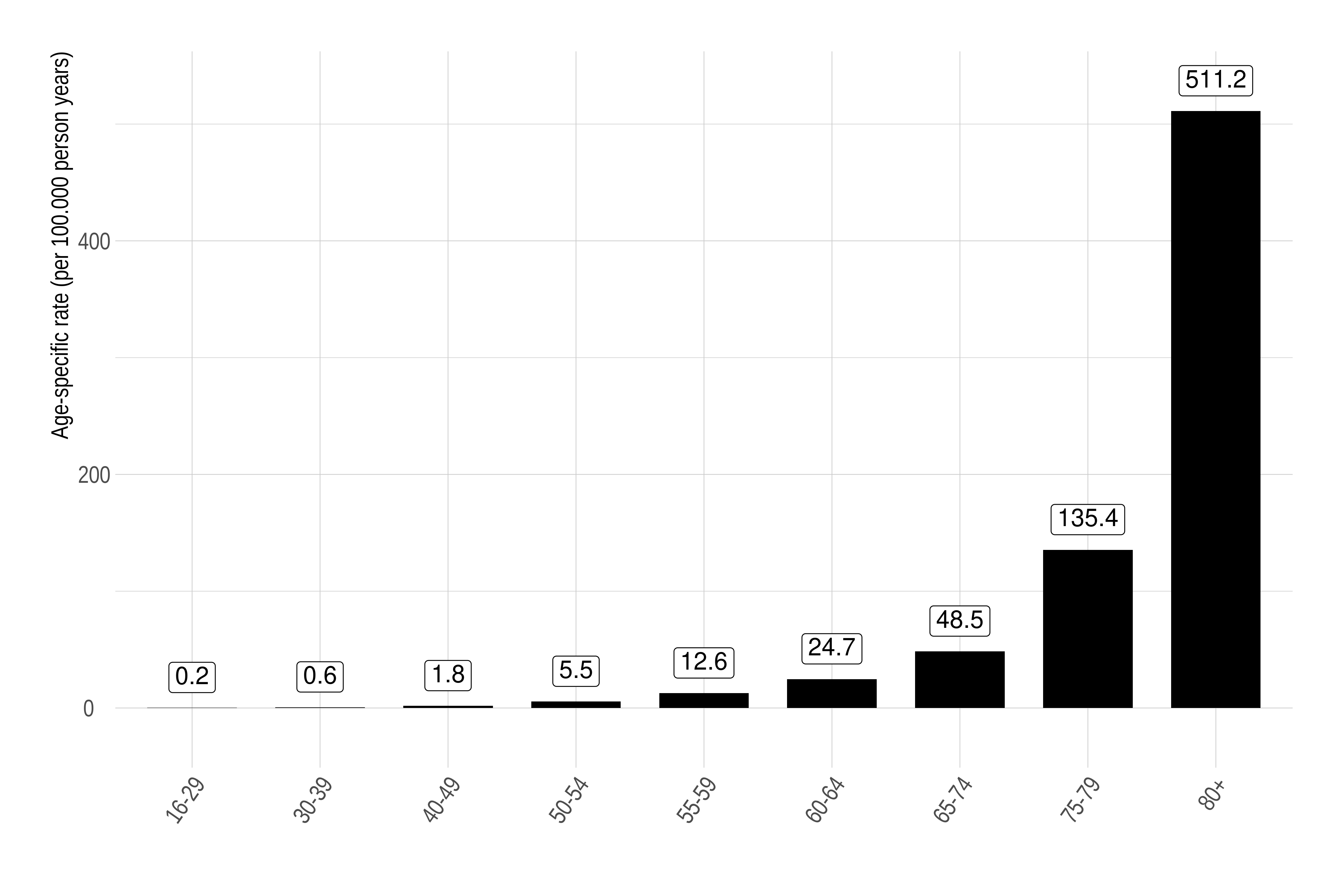

### Fig S2

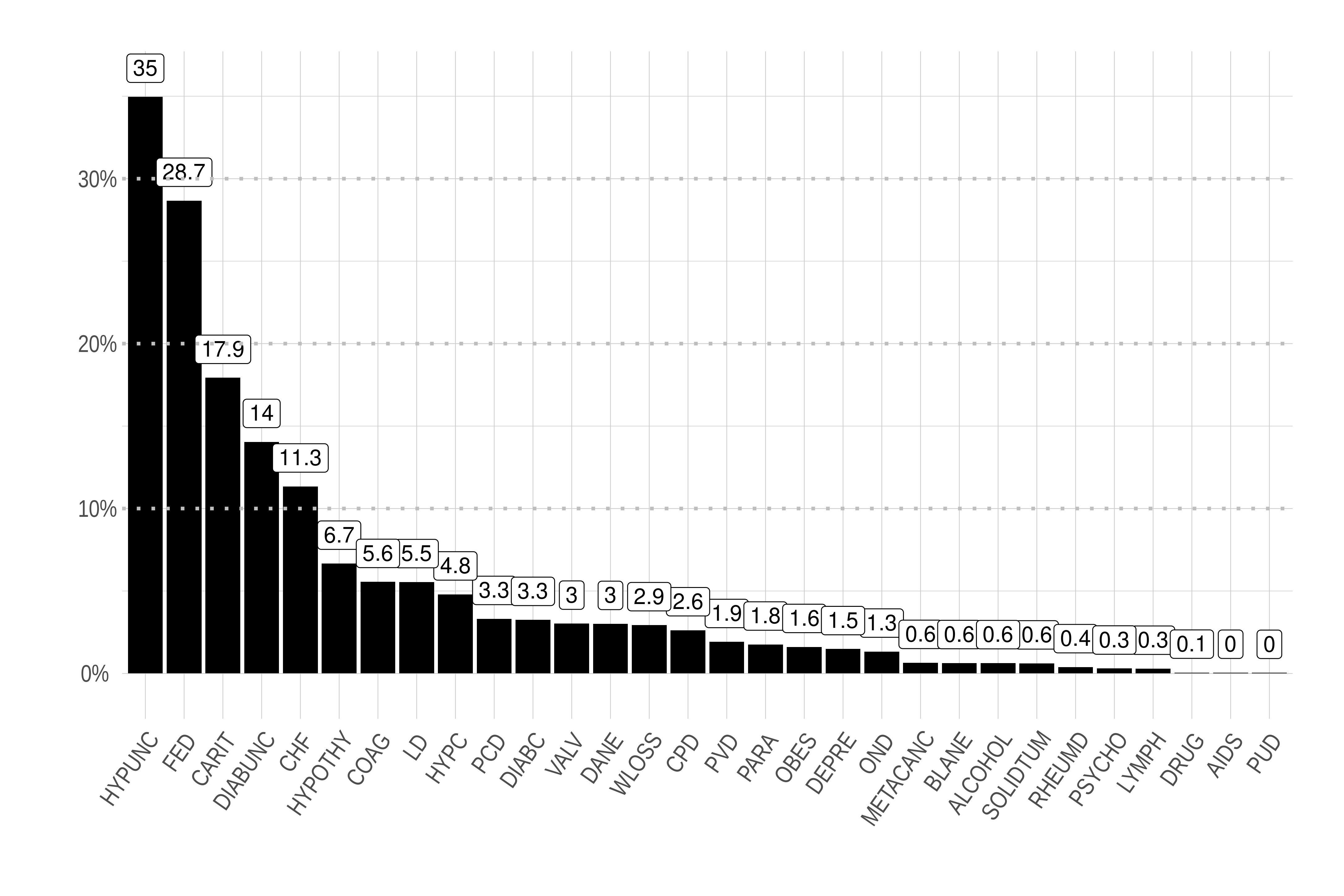

### Fig S3

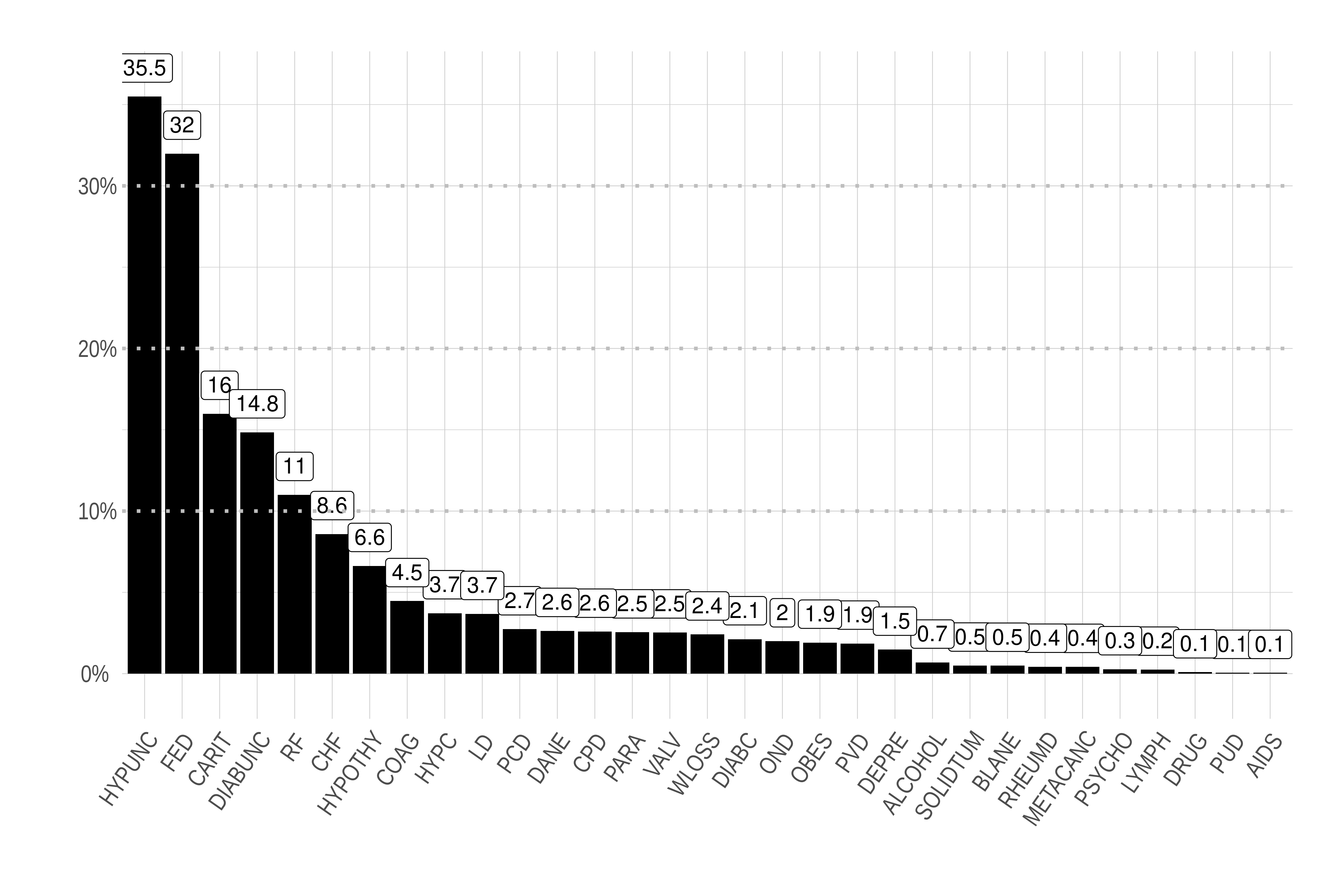

### Fig S4

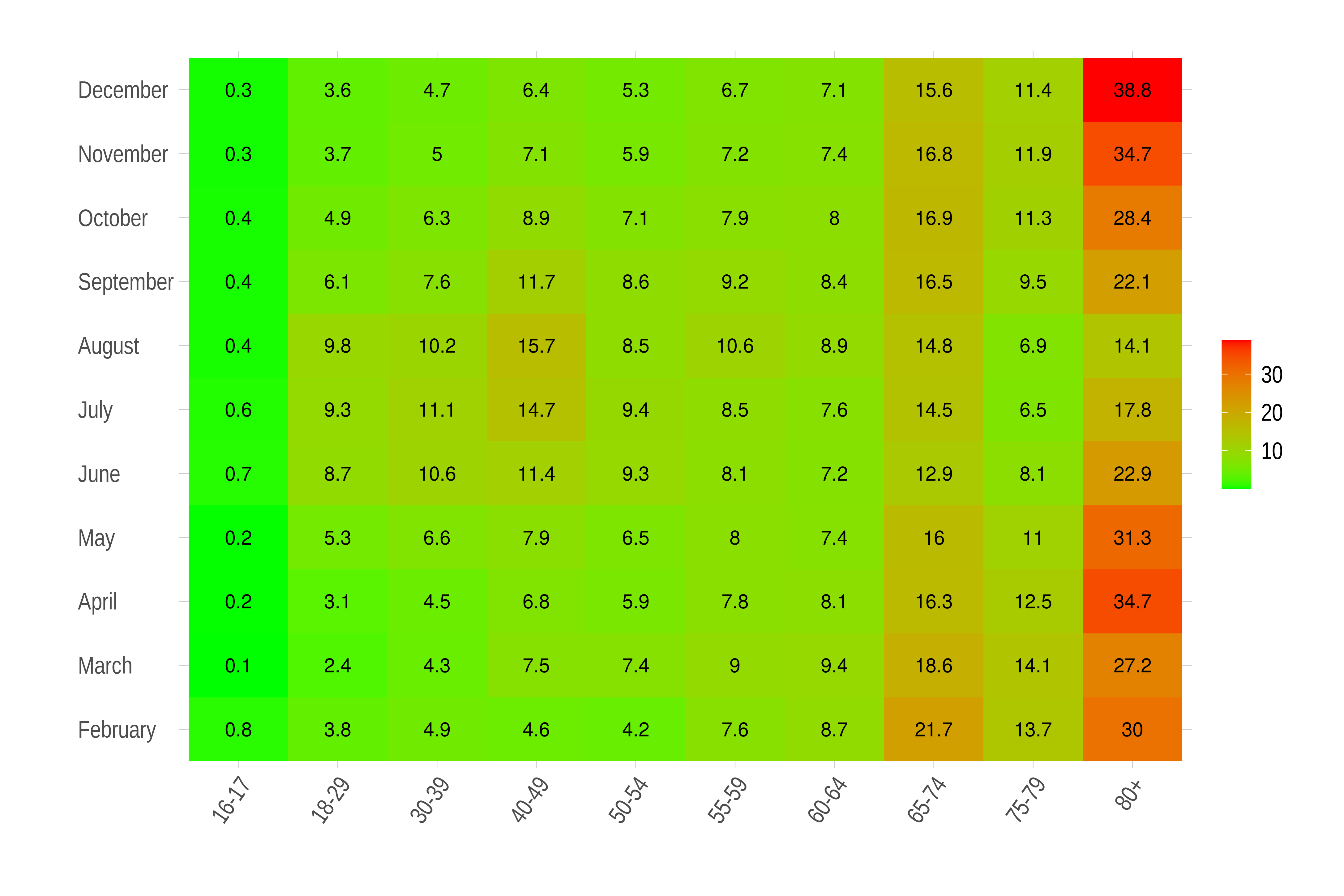

### Fig S5

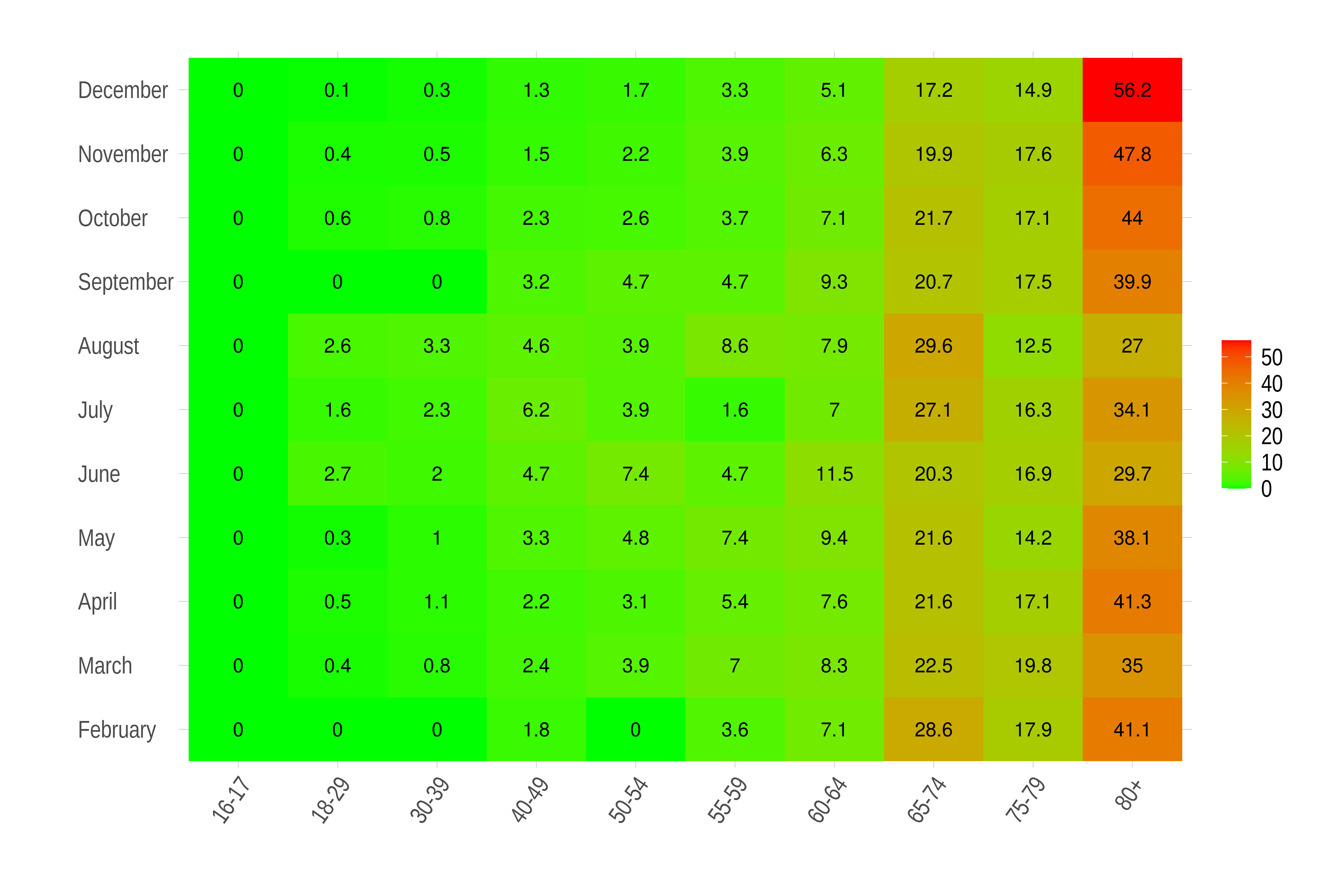
